# The collective wisdom in the COVID-19 research: comparison and synthesis of epidemiological parameter estimates in preprints and peer-reviewed articles

**DOI:** 10.1101/2020.07.22.20160291

**Authors:** Yuejiao Wang, Zhidong Cao, Daniel Dajun Zeng, Qingpeng Zhang, Tianyi Luo

## Abstract

**Summary:** *Background:* Research papers related to COVID-19 have exploded. We aimed to explore the academic value of preprints through comparing with peer-reviewed publications, and synthesize the parameter estimates of the two kinds of literature.

*Method:* We collected papers regarding the estimation of four key epidemiological parameters of the COVID-19 in China: the basic reproduction number (R_0_), incubation period, infectious period, and case-fatality-rate (CFR). PubMed, Google Scholar, medRxiv, bioRxiv, arRxiv, and SSRN were searched by 20 March, 2020. Distributions of parameters and timeliness of preprints and peer-reviewed papers were compared. Further, four parameters were synthesized by bootstrap, and their validity was verified by susceptible-exposed-infectious-recovered-dead-cumulative (SEIRDC) model based on the context of China.

*Findings:* 106 papers were included for analysis. The distributions of four parameters in two literature groups were close, despite that the timeliness of preprints was better. Four parameter estimates changed over time. Synthesized estimates of R_0_ (3·18, 95% CI 2·85-3·53), incubation period (5·44 days, 95% CI 4·98-5·99), infectious period (6·25 days, 95% CI 5·09-7·51), and CFR (4·51%, 95% CI 3·41%-6·29%) were obtained from the whole parameters space, all with p<0·05. Their validity was evaluated by simulated cumulative cases of SEIRDC model, which matched well with the onset cases in China.

*Interpretation:* Preprints could reflect the changes of epidemic situation sensitively, and their academic value shouldn’t be neglected. Synthesized results of literatures could reduce the uncertainty and be used for epidemic decision making.

*Funding:* The National Natural Science Foundation of China and Beijing Municipal Natural Science Foundation.

**Research in context:** *Evidence before this study:* Since its outbreak, scientific articles about the COVID-19 have greatly surged, with a significant portion as non-peer-reviewed preprints. Although preprints captured great attention, the credibility of preprints was widely debated. We searched PubMed and Google on March 20, 2020, for publications that discussed the preprints during the COVID-19 pandemic, using the terms (“preprints” AND “COVID-19”). We identified 12 papers and news, and found that scientists were skeptical of preprints mainly because rigorous peer review is absent and thus the conclusions of preprints may not be reliable. However, scientists’ opinions could have been biased towards limited data, and there is few knowledges about the validity of the results reported in the preprints. Further, to examine how scientists utilize results of preprints, taking the epidemiological parameter estimation as the objects, we searched reviews on Google using the terms (“epidemiology” AND (“meta-analysis” OR “reviews”) AND “COVID-19”) on May 23, 2020. Nine papers were identified. We found that existing meta-analysis and reviews included few preprints. This may be due to the fact that the quality of preprints was not recognized, and thus their academic value was underestimated. Overall, the validity of the results as reported in the preprints should be further examined and the potential of synthesizing preprints with formally published papers should be explored.

*Added value of this study:* Our study adds value in four main ways. First, we collected preprints and peer-reviewed papers on estimations of the four most important epidemiological parameters (the basic reproduction number, incubation period, infectious period, and case-fatality-rate) for the COVID-19 outbreak in China. 106 papers were included and available data were extracted. Second, we quantitatively compared the differences and timeliness between preprints and peer-reviewed publications in the estimation of the four parameters, and found that the validity of the preprints’ estimations was largely consistent with that of the peer-reviewed group. Third, we synthesized the estimations of the two groups of literatures using bootstrap method, and found that the values of infectious period and case-fatality-rate decreased over time, indicating that the synthesized results timely reflected the changing trend of the COVID-19 in China. Finally, the practicability of the synthesized parameter estimations was verified by the data of confirmed cases in China. The cumulative infection curve simulated using synthesized parameters fitted the real data well.

*Implications of all the available evidence:* Results of our study indicate that the validity of the COVID-19 parameter estimations of the preprints is on par with that of peer-reviewed publications, and the preprints are relatively timelier. Further, the synthesized parameters of the two literature groups can effectively reduce the uncertainty and capture the patterns of epidemics. These results provide data-driven insights into the academic value of preprints, which have been arguably underestimated. The scientific community should actively capitalize the collective wisdom generated by the huge amount of preprints, particularly during the emerging infectious diseases like the COVID-19.

## Introduction

The outbreak of COVID-19 posed a significant global threat. In response to the emerging infectious disease, the number of research papers has exploded in both formal publications and preprints.^1^ Many journals had the fast track to publish COVID-19 research, and made all COVID-19 work freely accessible to facilitate the information sharing. In contrast to previous Zika and Ebola outbreaks, scientists were more enthusiastic about posting articles on preprint archives this time due to the very high transmissibility of the COVID-19.^2,3^ Many major results were first posted online as preprints before being formally published in journals. But there were also voices questioning preprints’ authority^4-6^, believing that preprints pose the risk of dissemination of unconfirmed results and even rumors as they were not peer reviewed. However, the validity of the preprints has not been fully examined. Currently, scientists are overwhelmed by mixed and sometimes contradictory conclusions^7^, and the scientific community and policymakers face new challenge: how valid are the results of the preprints compared to journal papers and how to comprehensively integrate results from massive studies efficiently?

There are tons of preprints and peer-reviewed articles that estimated the four epidemiological parameters: the basic reproduction number (R_0_),^8^ incubation period, infectious period, and case-fatality-rate (CFR). It’s critical to accurately estimate these four parameters, because they indicate the transmission dynamics and severity of COVID-19. Based on various cases data sets and methods, estimates of preprints and peer-reviewed papers were varying over time. Several studies had reviewed the epidemiological parameters estimates. Maimuna et al. discovered 11 studies related to R_0_ estimation on Google Scholar and four preprint servers by Feb 9, 2020.^9^ They used a consensus-based approach to yield average R_0_ estimates for preprints and journal papers. Minah et al. searched on PubMed and preprint archives on Feb 21, 2020 and listed all estimates of R_0_, incubation period, and CFR.^10^ This study didn’t analyze the differences between preprints and peer-reviewed papers, nor did it propose a reasonable method to synthesize various results. Alqahtani et al. searched MEDLINE and Google scholar from inception date to March 16, 2020 and didn’t include preprints in the formal analysis of severity.^11^ And other meta-analysis on epidemiology contained only a small number of preprints.^12-14^ The scientific value of the preprints was largely overlooked by most of the reviews. However, we argue that the collective wisdom contained in the large number of preprints shouldn’t be neglected. And the potential of synthesizing preprint results with journal paper results should be explored.

Taking epidemiological parameters as objects, we aimed to quantitatively compare the validity of the preprints with peer-reviewed papers, and to synthesize the estimations of the two types of literature to mitigate the impact of uncertainty. This study compared and synthesized results for four parameters estimates (R_0_, incubation period, infectious period, and CFR) in two literature groups and two pandemic stages. Further, based on the historical data of COVID-19 in China, we evaluated the effectiveness of the synthesized parameters in predicting the epidemic trend.^8^ Our findings explored the collective wisdom in an epidemic crisis and indicated the academic value of the preprints.

## Materials and Methods

### Search strategy and selection criteria

Because the COVID-19 outbreak in mainland China has been basically contained since end March, we only searched and analyzed papers about the epidemic in China. We searched PubMed, Google Scholar and four popular preprint servers (i.e. medRxiv, bioRxiv, arRxiv and SSRN) for papers published from 23 January to 20 March, 2020 using the following terms: “2019-nCoV”, “coronavirus” or “COVID-19”. Through fast title screening, we removed papers focused on clinical treatment or papers whose research scopes were other countries instead of China. Then, the full-text screening was operated to remove comments, news, or papers that didn’t contain estimates for any of the following epidemiological parameters: (i) R_0_, the average number of secondary cases generated by an index case in the totally susceptible population; (ii) incubation period, the average time from infection to illness; (iii) infectious period, the period of time when an infected person is capable of transmitting the virus to others; (iv) CFR, the percentage of patients who die from a given disease. Finally, we noted that some of the later published papers directly adopted the earlier estimate. Among papers related to the incubation period, 12 cited the same paper published in New England journal of Medicine on January 29,^15^ which estimated the incubation period to be 5·2 days. So, we removed the papers that adopted the same estimate of an earlier paper. For the preprint that have been published in a certain journal by 20 March, we only keep the journal version.

### Data analysis

The following information was manually extracted from each paper: title, publication date (T_P_), manuscript submission date (T_S_), publication source, estimates for the corresponding four parameters (R_0_, incubation period, infectious period, and CFR), and their uncertainty intervals (if available). The publication delay (T_D_) of each paper was calculated by the difference between T_P_ and T_S_. Since a few peer-reviewed papers didn’t provide T_S_, the latest date for cases data collection in that paper was approximated as T_S_. Based on publication sources, the literature collection was divided into preprints group and peer-reviewed group.

To compare the parameter estimations and timeliness between the preprints and peer-reviewed papers, the distributions of four parameters estimates and T_D_ of the two groups were separately plotted using the “seaborn” toolbox in Python 3·7·3. Next, we used the bootstrap method to estimate the means and 95% confidence intervals (95% CIs) of the four parameters in the two groups.^16^ Means of the entire parameter set (not grouped) were obtained as well. The bootstrap method was conducted by the built-in function “bootci” of Matlab R2017a.

With more awareness of the COVID-19 outbreak and more data being accumulated, the parameter estimates changed over time. To integrate results from literatures in the time dimension, the estimations of R_0_, incubation period, infectious period, and CFR were ranked in the chronological order based on T_P_. Taking February 13 as the demarcation point, the whole period from January 23 to March 20 were divided into two stages. Stage one was from January 23 to February 12, and stage two was from February 13 to March 20. The first stage was the epidemic development period, and the second stage was the epidemic recession period. Because the Chinese government isolated and treated 14,840 mild or clinically confirmed cases in Hubei Province on February 12, which further prevented the interpersonal transmission of the virus, the number of daily confirmed cases in China began to decline on February 13. The same bootstrap method was harnessed to calculate the iterative updates of parameter estimates over time and the means of the four parameters in the two stages, respectively.

To evaluate the effectiveness of the four synthesized parameters in predicting the epidemic trend, we randomly assembled the means of bootstrap samples of the four parameters of the whole dataset and put them into the SEIRDC model (supplementary material, p. 6-7).^8^ Considering the outbreak of COVID-19 in mainland China, 1·4 billion population in China were set as the susceptible population with an initial infected person. After 1000 Monte Carlo simulations of SEIRDC model, we obtained the mean curves of simulated cumulative infected cases (C(t)). On the other hand, the cumulative onset infections in China after epidemiological retrospective investigation were obtained from the report of the WHO-China joint mission on Coronavirus Disease 2019. The simulated curve and real curve were aligned and compared based on the date when the cumulative infections reached 100. Accumulated infections of China by March 20 were also obtained from the official website of National Health Commission of China.

## Results

After selection from the 339 potentially literatures (figure 1), 106 papers were included in the final collection. 58 (54·7%) papers contained estimates for more than one parameter. There were far more preprints than peer-reviewed papers: preprints accounted for 78·0% (32/41), 72·3% (34/47), 79·3% (23/29), and 84·6% (22/26) of the papers related to R_0_, incubation period, infectious period, and CFR, respectively. Papers included and the main characteristics (four parameters, T_P_, T_S_, and T_D_) extracted from them were summarized (supplementary table 1-4, p. 1-5).

**Figure 1.**
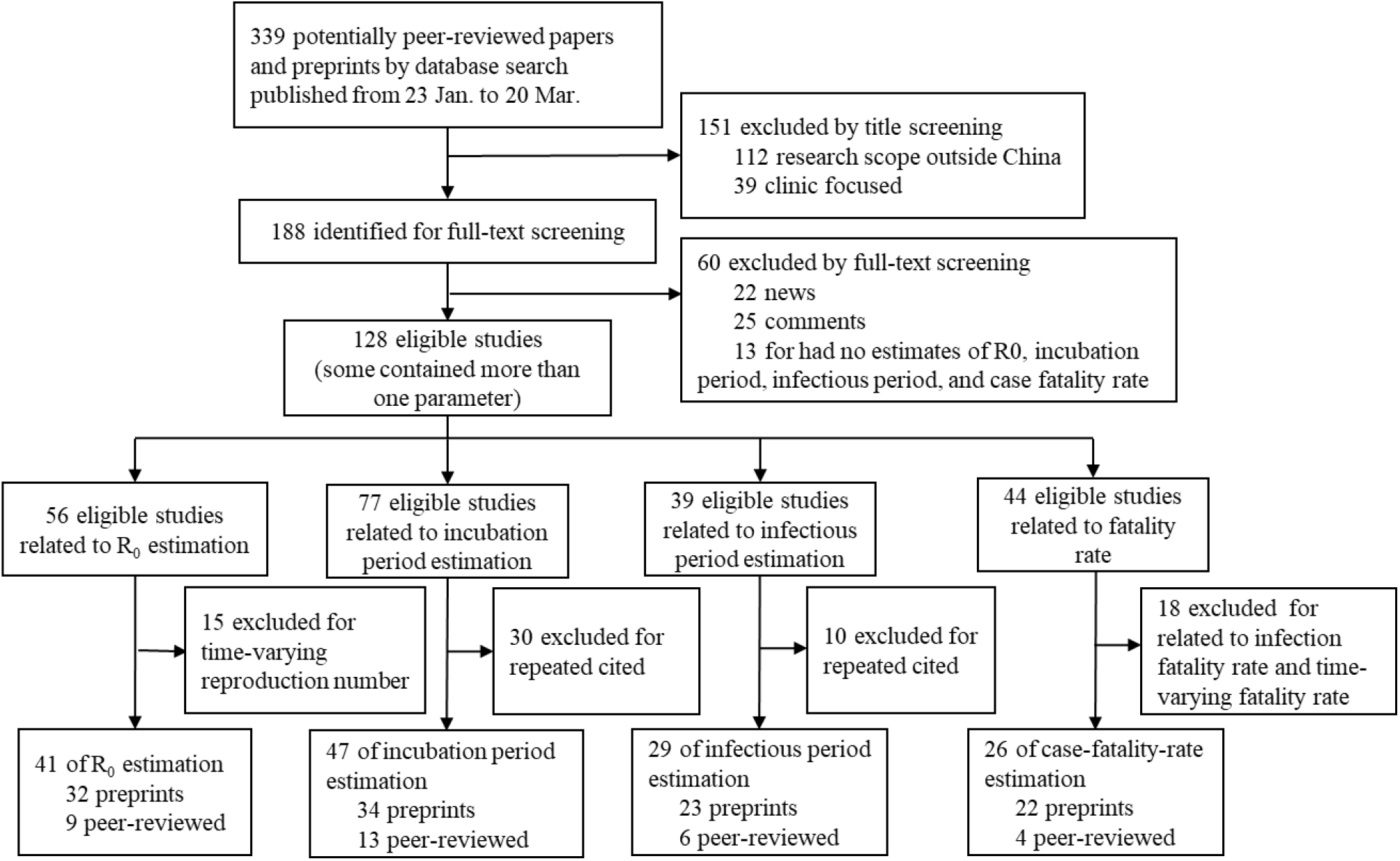
Study selection process

The distributions and quantiles of R_0_, incubation period, and infectious period in two groups were close (figure 2a), but the quartiles of CFR in preprints group (2·84%, IQRs 1·38%-5·13%, p<0·05, figure 2a) was much lower than that of in peer-reviewed group (5·6%, IQRs 4·7%-8·1%, p<0·05). As for the comparison of timeliness, T_D_ of preprints were much lower than peer-reviewed papers (figure 2b). The quantiles of T_D_ of preprints were 1 (IQRs 0-2, p<0·05), 2 (IQRs 1-3·25, p<0·05), 2 (IQRs 1-2·25, p<0·05), and 2 (IQRs 1-3, p<0·05), corresponding to the four parameters, respectively. While the review speed of different journals varied greatly: the T_D_ of peer-reviewed papers were 7 (IQRs 5-16, p<0·05), 7 (IQRs 2-24·5, p<0·05), 5 (IQRs 2-31, p<0·05), and 16 (IQRs 10-16·75, p<0·05), respectively.

**Figure 2.**
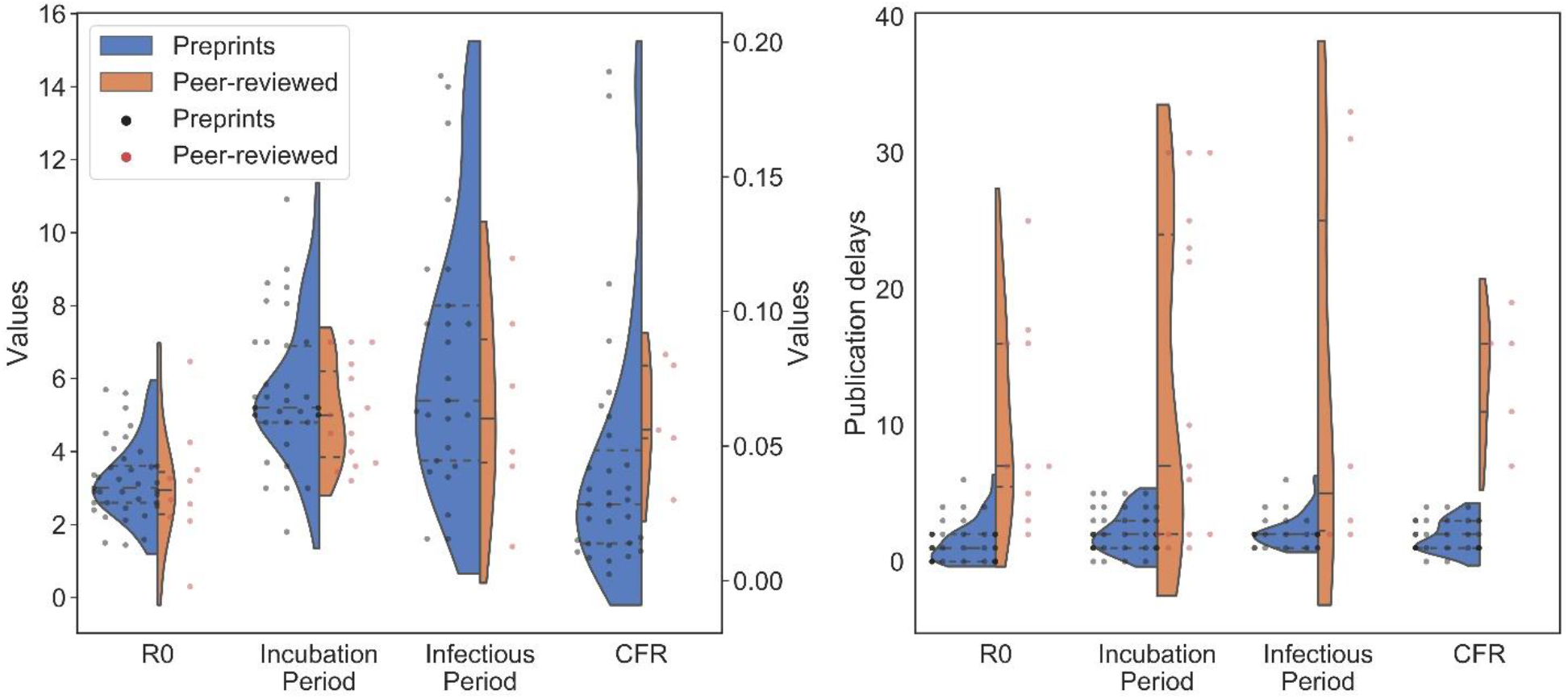
(a) Full distributions of estimates of R_0_, incubation period, infectious period, and CFR and (b) their corresponding publication delays in preprints groups and peer-reviewed group. CFR corresponds to the right y-axis in figure 2a.

Synthesized estimations of the four parameters of each literature group generated by bootstrap method were listed in table 1. We estimated the mean R_0_ in preprints group to be 3·20 (95% CI 2·92-3·59, p<0·05), slightly higher than mean R_0_ of 3·07 (95% CI 2·23-4·17, p<0·05) in peer-reviewed group. Similarly, the mean incubation period (5·61 days, 95% CI 5·07-6·29, p<0·05) and infectious period (6·54 days, 95% CI 5·24-8·08, p<0·05) in preprints group, were still longer than that in peer-reviewed group, which were 5·04 (95% CI 4·41-5·72, p<0·05) days and 5·25 (95% CI 3·32-7·25, p<0·05) days. Estimate of CFR in preprints group was 4·26% (95% CI 3·10%-6·31%, p<0·05), but the mean CFR in peer-reviewed group (6·10%, 95% CI 4·00%-7·62%, p<0·05) was much higher and with a smaller range of uncertainty.

**Table 1.** Estimates of the basic reproduction number (R_0_), incubation period, infectious period, and case-fatality-rate (CFR) generated by bootstrap method.

| Groups | $R_0$ | Incubation period<br>(days) | Infectious period<br>(days) | CFR (%) |
| --- | --- | --- | --- | --- |
| **Preprints** | 3.20<br>(95% CI 2.92-3.59) | 5.61<br>(95% CI 5.07-6.29) | 6.54<br>(95% CI 5.24-8.08) | 4.26<br>(95% CI 3.10-6.31) |
| **Peer-reviewed** | 3.07<br>(95% CI 2.23-4.17) | 5.04<br>(95% CI 4.41-5.72) | 5.25<br>(95% CI 3.32-7.25) | 6.10<br>(95% CI 4.00-7.62) |
| **Stage One** | 3.10<br>(95% CI 2.64-3.73) | 5.14<br>(95% CI 4.63-5.63) | 7.19<br>(95% CI 4.86-9.65) | 5.93<br>(95% CI 3.27-11.42) |
| **Stage Two** | 3.25<br>(95% CI 2.89-3.64) | 5.63<br>(95% CI 5.00-6.50) | 5.76<br>(95% CI 4.73-7.38) | 4.09<br>(95% CI 3.09-6.24) |
| **Overall** | 3.18<br>(95% CI 2.85-3.53) | 5.44<br>(95% CI 4.98-5.99) | 6.25<br>(95% CI 5.09-7.51) | 4.51<br>(95% CI 3.41-6.29) |

\*all 95% CI with $p < 0.05$ .

Regardless of groups, iterative updates of four parameters estimates in time dimension were shown in figure 3, and the synthesized estimations for the two pandemic stages were shown in table 1. Except that the CFR data points in stage one were significantly less than those in stage two, the data of the other three parameters was relatively evenly distributed (figure 3). The mean R_0_ in stage one (3·10, 95% CI 2·64-3·73, p<0·05) was close with R_0_ in stage two (3·25, 95% CI 2·89-3·64, p<0·05). The mean incubation period (5·14 days, 95% CI 4·63-5·63, p<0·05) in stage one was slightly shorter than that in stage two (table 1). But the mean infectious period declined from 7·19 (95% CI 4·86-9·65, p<0·05) days to 5·76 (95% CI 4·73-7·38, p<0·05) days. Because of limited data, mean CFR in stage one was 5·93% with a larger 95% CI between 3·27% and 11·42%, and CFR in stage two was 4·09% (95% CI 3·09%-6·24%, p<0·05). The overall estimations of these four parameters were also given in table 1. The gaps between the overall estimates and the estimates of the preprint group were smaller, because preprints accounted for the majority.

**Figure 3.**
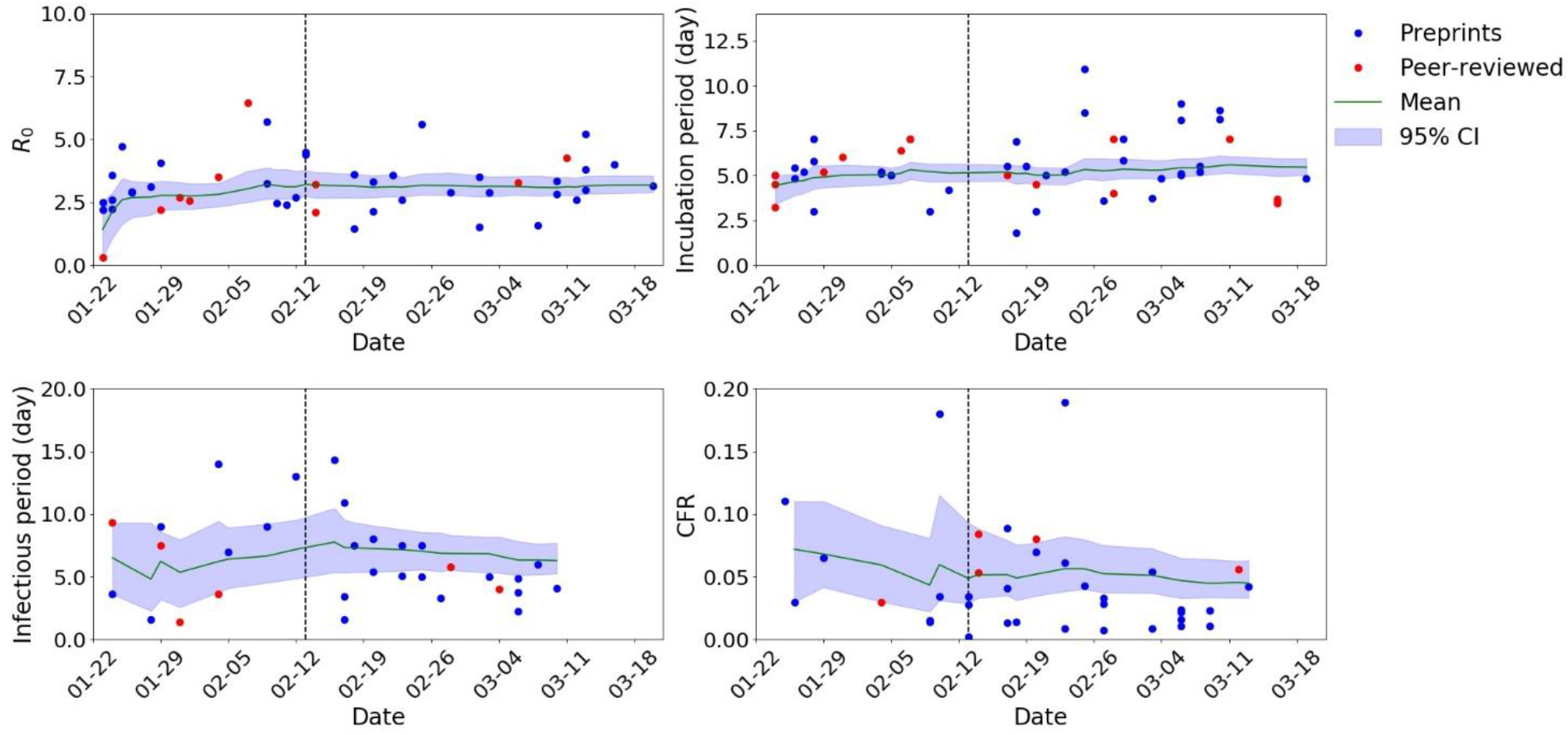
Distributions of R_0_, incubation period, infectious period, and CFR ordered by publication time.

To evaluate effectiveness of the four synthesized parameters in predicting the epidemic trend, C(t) of mainland China from December 26, 2019 to February 19, 2020, obtained by 1000 Monte Carlo simulation of SEIRDC model, were shown in figures 4 (please refer to the method section for the specific simulation method). December 26 was the date when both the cumulative onset infections (the blue curve) and simulated infections (the red curve) reached 100. By December 31, 2019, the cumulative onset cases almost exactly matched the simulated mean (figure 4). From January 1 to February 7, the onset infections were slightly above the simulated mean, but still within the range of simulations. During this period, China has taken many prevention and control measures, including traffic restriction and makeshift hospitals in Wuhan. After February 7, the epidemic in China was effectively contained by quarantine and treatment measures, and as of February 19, the cumulative onset infections of China had stabilized at about 75,100 (figure 4). The officially reported cumulative confirmed cases (the green curve) significantly lagged the onset infections in the early period, and the two curves were not roughly equal until February 13.

**Figure 4.**
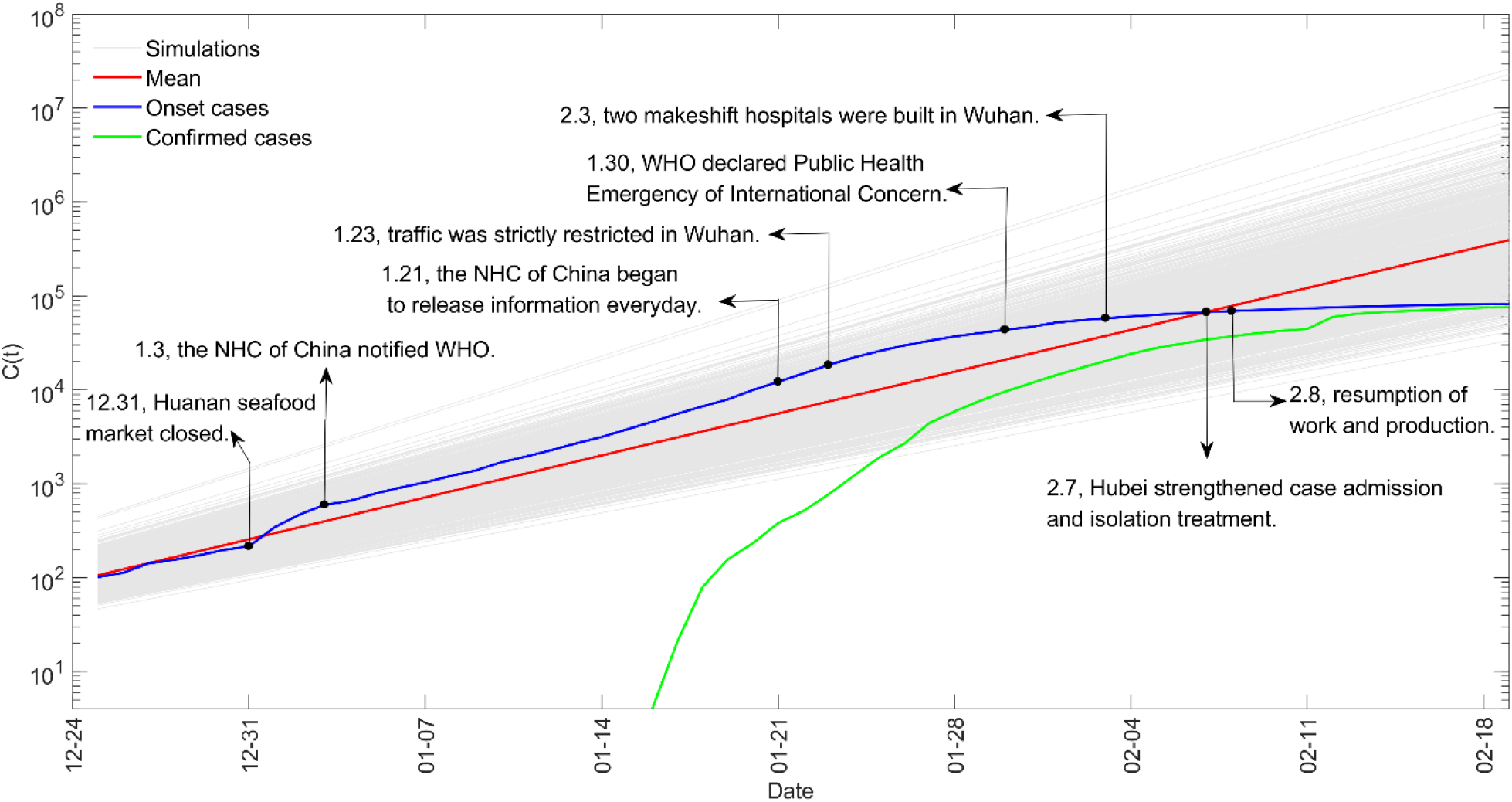
The simulation results of cumulative number of infections of China.

## Discussion

In this study, based on literatures of COVID-19 in mainland China by March 20, 2020, we compared distributions of parameter estimations between preprints group and peer-reviewed group, and synthesis the estimations according to groups or publication date. Results showed that, except for CFR, the distributions and synthesized estimates of R_0_, incubation period, and infectious period were similar between the two groups. Estimates of R_0_ and incubation period remained stable in two pandemic stages, but the estimates of infectious period and CFR in stage two declined significantly. Further, the SEIRDC simulations of COVID-19 outbreak in China evaluated the applicability and validity of the comprehensive parameters space. The actual cumulative onset infections and the simulation results matched well.

The validity of the preprints in parameter estimation was quantitatively analyzed in this study. The validity of preprints was always controversial, and there were far more preprints than peer-reviewed papers because they were simply reviewed by volunteers on the platform. Due to the same reason, preprints are also much timelier. Some scientists argued that the conclusions of the preprint may be misleading and should not be widely adopted,^17^ while others thought that because the preprint was available to the public, authors would pay more attention to their personal reputations, and the quality of the preprint would not be uncontrolled.^18,19^ The comparison results of this study showed that the estimated distributions of R_0_, incubation period and infectious period in the preprints group were similar to that of the peer-reviewed group (figure 2a). And the distributions of preprints were more concentrated, so the ranges of 95% CIs of preprints were smaller (table 1). These suggested that, in the outbreak of COVID-19, although the result of individual preprint may be biased, the validity of synthesized parameter estimates of the preprints were at the same level as the peer-reviewed papers, and the synthesized estimations of preprints were even more robust. Therefore, it is not wise to neglect the collective wisdom contained in the large number of preprints.

The iterative estimations of parameters from the time dimension can reflect the trend of the epidemic. Compared with stage one, the corresponding infection period in stage two was shorter and CFR was reduced (table 1). The possible reason for the changes was that many patients diagnosed in the late period were included for the parameter estimation in studies that posted in stage two. Due to effective control measures in China, the speed of testing was improved, and the cure rate also increased, so the infectious period and CFR decrease. This indicated that integrating epidemiological parameters in the time dimension could also reflect changes of the epidemic situation. Besides, in this study, the preprints accounted for 72%-85% of the total literature, so these trends were largely reflected by preprints. And since the publication delay of preprint was shorter (figure 2b), the preprints allowed us to get the latest information to assist emergency decision-making in a timely manner.

The results of many papers inevitably have uncertainties. It is unreasonable to make decision based on the conclusion of a single paper. To mitigate the effects of uncertainty, it is more robust to synthesize the results from multiple papers. In this study, through random sampling and 1000 Monte Carlo simulations, the four parameter estimates of published papers were utilized in a comprehensive way. In figure 4, in the absence of human intervention, before the closure of the Huanan seafood market, the cumulative onset cases and the simulated mean were almost in line. After January 23, due to strong isolation and control measures, the rate of cumulative onset cases slowed down. The cumulative onset cases were always within the uncertain range of the simulations. However, at that time, the official confirmed cases (figure 4) that scholars could obtain for analysis were far behind the real onset cases. It indicated that even if scientific data were lacking and delayed in the early period of COVID-19, the whole parameter space still well grasped the pattern of epidemic spread. This not only reminds us to comprehensively refer to the results of all published papers, but also reflects the practical value of the preprints because they are the majority.

This study has some limitations. Firstly, the data officially reported by China didn’t fully represent all the infections and deaths. Because in the early period, many patients died without diagnoses. And with the huge burden on the medical system in Hubei Province, it was impossible to detect and report all cases without omission. We can only prove the validity of our parameter estimations to a certain extent, but we can’t deny the reference value of collective wisdom from literatures.

Secondly, in our study, the validity of the preprints was only compared and evaluated on the overall distribution, demonstrating their academic value on the task of estimating the four epidemiological parameters of COVID-19. However, this doesn’t mean that the result of a single preprint is accurate, nor can the conclusion of our study be arbitrarily extended to other fields. Scientists should treat preprints with caution and responsibility, and we should further standardize the publication process of preprints and guide the media to scientifically report preprints.

In conclusion, our quantitative analysis shows that the overall validity of the preprints in parameter estimation is not less than that of the peer-reviewed papers. And the latest information on the epidemic can be obtained more sensitively through preprints. Furthermore, the simulation of the COVID-19 in China proved that the synthesis of whole parameters space is an effective way to reduce the uncertainty and to grasp the pattern of transmission. In response to future public health crises, scientists should be more proactive in promoting the development of preprint platforms and quality monitoring,^2^ while more automated literature analysis and integration methods should be developed to make collective intelligence more applicable to decision making.

## Data Availability

The data that support the findings of this study are available from the supplemental material and it can be open accessed.

## Contributors

YW, ZC, DDZ, and QZ conceived the study. YW, ZC, and TL developed the methods. YW and TL collected the data. YW wrote the first draft of the manuscript and supplemental material. QZ and ZC provided critical feedback and contributed to the writing of the manuscript.

## Acknowledgments

This study was funded by National Natural Science Foundation of China (Nos. 72042018,91546112, 71621002) and Beijing Municipal Natural Science Foundation (No. L192012).

